# Patient information needs for transparent and trustworthy artificial intelligence in healthcare

**DOI:** 10.1101/2024.07.02.24309850

**Authors:** Austin M. Stroud, Sarah A. Minteer, Xuan Zhu, Jennifer L. Ridgeway, Jennifer E. Miller, Barbara A. Barry

## Abstract

**Background:** As health systems incorporate artificial intelligence (AI) into various aspects of patient care, there is growing interest in understanding how to ensure transparent and trustworthy implementation. However, little attention has been given to what information patients need about these technologies to promote transparency of their use.

**Methods:** We conducted three asynchronous online focus groups with 42 patients across the United States discussing perspectives on their information needs for trust and uptake of AI, focusing on its use in cardiovascular care. Data were analyzed using a rapid content analysis approach.

**Results:** Our results suggest that patients have a set of core information needs, including specific information factors pertaining to the AI model, oversight, and healthcare experience, that are relevant to calibrating trust as well as perspectives concerning information delivery, disclosure, consent, and physician AI use.

**Conclusions:** Identifying patient information needs is a critical starting point for calibrating trust in healthcare AI systems and designing strategies for information delivery. These findings highlight the importance of patient-centered engagement when considering approaches for transparent healthcare AI.

## Introduction

A chief concern among published ethical frameworks and guidance documents for responsible healthcare artificial intelligence (AI) is the lack of transparency regarding how AI models—computer software trained to recognize patterns in health data—have been developed and trained.^1-7^ These models are often opaque without insight into how or why a particular output was derived, raising concerns about transparency even if the output proves to be measurably accurate.^8^ This “black-box” problem poses a unique challenge in healthcare settings, where well-informed and transparent clinical decision-making is critical to healthcare delivery and patient trust.^9^ There are also implications for health equity, with greater transparency being a potential mechanism for mitigating bias in models.^10^ Furthermore, physicians may need to grapple with appropriate disclosure of AI use and associated risks, particularly when details about model development and training are lacking; nonetheless, concerns exist that insufficient AI transparency may be disruptive to physician-patient relationships which are essential for determining proper clinical investigation and treatment in response to AI outputs.^11-13^

Efforts to facilitate transparency for healthcare AI tools include novel technical approaches to illuminating model development processes and creating more explainable AI (XAI) tools.^14, 15^ For example, visualizations for deep learning systems in cardiovascular disease modeling aim to support understanding by highlighting interactions between variables.^16^ However, these technical approaches are limited and XAI only serves as one mode for enhancing transparency in a system with numerous stakeholders of varying information needs, particularly patients who may lack the technical skills and training to interpret these outputs.^17^

In addition to technical approaches, there is growing scholarship around AI model documentation—the provision of information accompanying AI tools for enhanced transparency.^18, 19^ These approaches provide supplemental information and evaluative criteria about a given model or data set and, in some instances, can be dynamically tailored for different use cases or revised as models are updated.

Prototypes such as “AI nutrition labels” that display information similar to nutrition or prescription drug labels have been developed for technical and clinical end-users (i.e., software engineers, data scientists, and physicians). However, there is still a lack of consensus regarding the optimal approach for including and assessing information about healthcare AI, and these efforts have largely focused on enhancing transparency for a technical and clinical expert audience.^20-22^

There is limited empirical work focusing on enhancing transparency of AI in healthcare for patients. Recent qualitative research exploring AI information has examined the needs of patients regarding diabetes treatment, including creating tutorials for information access and alerts to guide device use.^23^ Other research assessing patient perspectives on AI in healthcare more broadly, or for specific tools, has identified patient concerns about transparency, including the potential for models to make unexplainable errors or limit patient decision-making.^24-26^ While current scholarship emphasizes transparency as a key principle, there is limited evidence on patients’ specific information needs. This knowledge gap creates a significant barrier to developing inclusive and transparent solutions and enhancing understanding of information factors—discrete elements of information communicated to a patient (e.g., model performance, manufacturer)—that influence patient trust in healthcare AI. There are also downstream implications for safety and efficacy, as model transparency may factor into clinician and patient decisions about AI use and related clinical recommendations.^27^

The present online focus group study aimed to identify patients’ core information needs if AI tools were to be used in their health care. To elicit patient perspectives, we used clinical vignettes depicting AI in the diagnosis, treatment, and/or monitoring of cardiovascular conditions and posed questions about healthcare AI in general. We also explored information factors that influence patient trust in these technologies, their preferences for information delivery, and their perspectives about physicians using AI in their care. Our overall objective is to provide a more robust evidentiary foundation for designing patient-centered transparency solutions for healthcare AI and calibrating trust in these tools.

## Methods

### Study Design

We conducted asynchronous online focus groups with patients across the United States to identify their information needs and preferences to consider transparent and trustworthy AI. Clinical vignettes depicting AI technologies in cardiovascular care were used as prompts for discussions along with general questions about healthcare AI broadly. This study is part of a larger multi-method investigation of clinician and patient information needs and ethical considerations for adoption of AI in healthcare.

### Ethics

Review and approval for this study and all procedures was obtained from the Mayo Clinic Institutional Review Board (protocol # 21-012302). Consent was obtained from participants verbally prior to each focus group in accordance with IRB guidance.

### Recruitment

Potential research participants were sampled via ResearchMatch, a national health volunteer registry with over 140,000 subjects who have agreed to be contacted about research opportunities. Several academic institutions created this database; it is supported by the U.S. National Institutes of Health as part of the Clinical Translational Science Award (CTSA) program.

We contacted participants via the ResearchMatch platform with a message containing a brief description of our study, eligibility criteria, and voting buttons that let subjects affirm or deny their interest. ResearchMatch provided contact information (e.g., name, email address) for those who responded affirmatively to the message. These eligible participants were added to our recruitment list. Participants were considered eligible if they were at least 18 years of age, English-literate, and had a primary care or cardiology clinical visit within the past three years. We oversampled from American Indian, Asian, Black/African-American, Native Hawaiian/Pacific Islander, Hispanic/Latino, and Multi-racial populations to increase their representation in our sample, as these groups have been underrepresented in US medical research.^28, 29^

### Data Collection

Three asynchronous online focus groups were conducted.^30^ Each focus group took place over two days on FocusGroupIt (focusgroupit.com). Focus groups were limited in size to up to 15 participants based on best practices that balances the depth and richness of discussion and participant and moderator burden.^30, 31^ Questions were pre-loaded into the FocusGroupIt platform by a member of the research team (AMS). The first set of questions were posted at midnight, and the second set of questions were posted 24 hours later. Participants responded to both free-response and multiple-choice questions to enhance the breadth of collected data. Some questions were presented alongside a hypothetical clinical vignette describing AI used in the care of a cardiovascular condition; a vignette example is depicted in Box 1. These vignettes were reviewed by subject matter experts for clinical accuracy and patient-centered communication.^32^ Participants reviewed four total vignettes adapted from cardiovascular technologies in industry and academic literature. Technologies depicted included 1) a smartwatch that monitored ECG readings, 2) a stethoscope that classified heart murmurs, 3) a monitoring tool that assessed a patient’s condition after a heart attack, and 4) an inpatient monitoring tool that assessed post-surgical sepsis risk.^33-36^ Participants were encouraged to log in on two consecutive days to answer the posted questions and comment on other participants’ posted responses. All participants were given an anonymous account name by FocusGroupIt when they logged in (i.e., Participant 1, Participant 2). On both days, four members of the research team (AMS, BAB, SAM, and XZ) reviewed participants’ posts throughout the day and posted follow-up questions and responses.

#### Box 1. Vignette example depicting healthcare AI used in online focus groups.

A patient comes to the emergency department with chest pain and shortness of breath. Dr. Rodriguez, an emergency medicine intern in training, takes a full history and performs a physical exam of the patient’s heart and lungs. During her examination, Dr. Rodriguez uses an artificial intelligence enabled stethoscope that provides enhanced listening of the patient’s heart and creates a recording for the physician to review after the examination. The artificial intelligence can detect nuances in heart sounds that may go unnoticed by inexperienced ears. After completing her examination, Dr. Rodriguez identifies a preliminary diagnosis of a heart murmur, sees the stethoscope’s readings also indicate that a heart murmur is likely, and schedules cardiology appointment for the patient to follow-up.

After participants responded to all questions, they were redirected to a separate survey hosted in REDCap where they could provide their contact information to receive $25 in remuneration.^37, 38^ Personal information entered in REDCap could not be linked to participants’ focus group responses. Participants who had agreed to participate in the focus group but had not entered information into REDCap at the end of day two, received an email reminder that the final focus group section provided a link for entering their information for remuneration. We left focus groups open for three days in case some participants needed extra time to complete their responses. Moderators did not post any follow-up prompts after the second day.

### Analysis

We applied a rapid analytic approach to the qualitative focus group data.^39^ All free-text responses from each focus group were exported verbatim and used to create individual summary tables that synthesized content according to thematic headings that corresponded to questions posed in the group. Three study team members conducted content analyses (AMS, SAM, and XZ).^40^ One team member generated initial summary tables, while another reviewed the tables for accuracy and supplemented content. Once completed, the summary tables were collapsed across focus groups and memoranda were generated that identified major themes. The study team discussed major themes and key findings and arrived at consensus. A thematic map illustrating patient information needs was developed based on an inductive analysis of participant responses to questions focused on AI transparency (i.e., *what would you want to know if AI was being used in your healthcare?*). Trust factors were also identified based on patient responses to questions focused on trust (i.e., *what would you need to trust the AI results or recommendations?)* as well as other responses directly referencing trust. Multiple choice question responses were aggregated, and descriptive statistics were generated using R (version 4.2.1).

## Results

Eighty-five participants were contacted via email with an invitation to participate in the study. Participants who responded (n=42) indicating an interest in participating were enrolled in a focus group. We conducted three focus groups with a total of 42 participants between October and November of 2022. Demographic characteristics of study participants are presented in Table 1. We report on main and sub-themes related to 1) information about AI development, oversight, and impact on care as well as trust factors, 2) preference for model documentation and information delivery via labels and other mechanisms, 3) importance of disclosure of AI use when critical to provider decision making or care provision, 4) patient perceptions of consent to use AI, 5) patient preferences for deciding whether AI is used in their care, and 6) favorable perceptions of physicians using AI. In addition to these findings, we present descriptive statistics for responses to attitudinal questions concerning AI transparency detailed in Table 2.

**Table 1:**
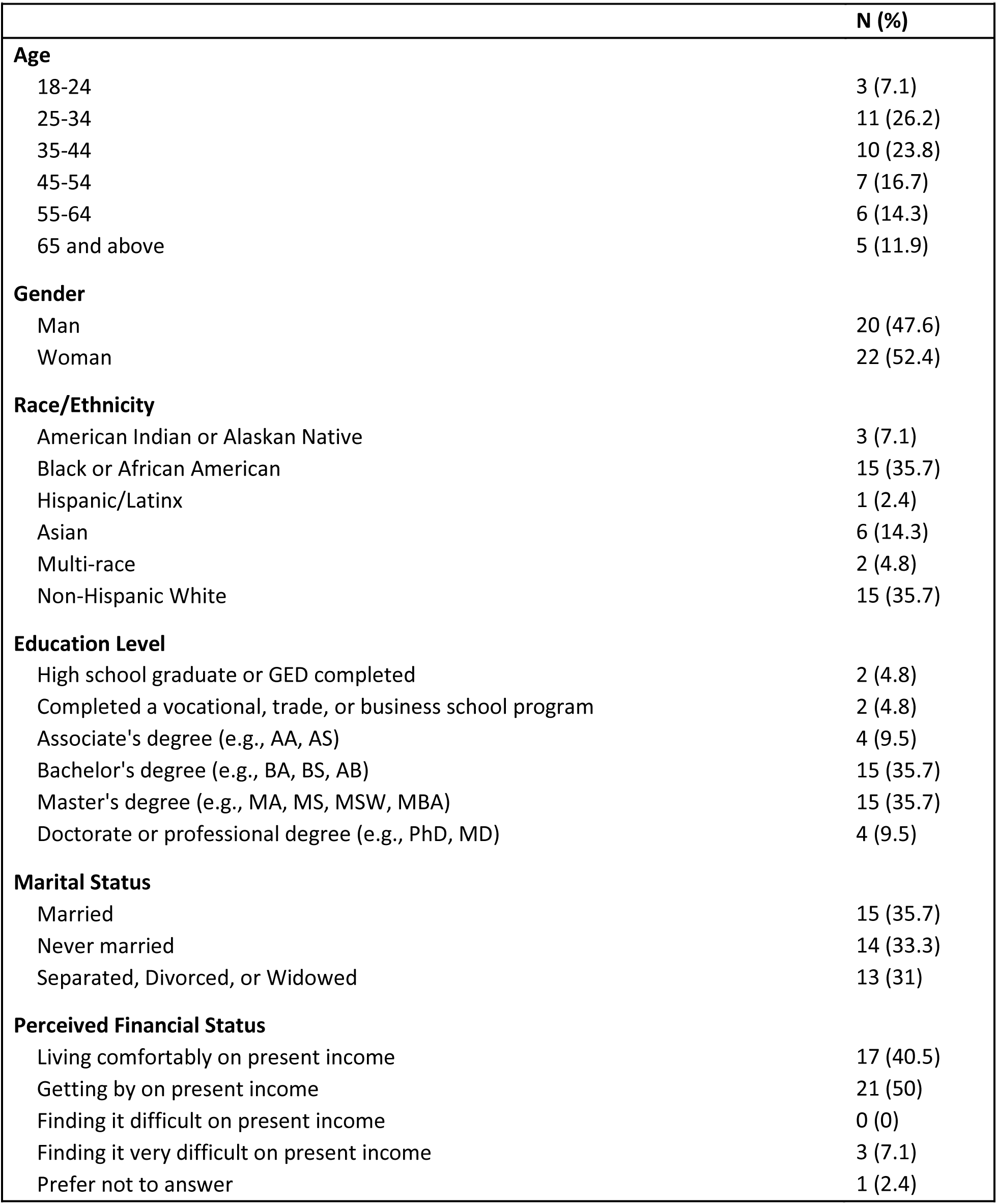
Demographic characteristics of 42 patients who participated in focus groups.

**Table 2:** Descriptive statistics of participant responses to attitudinal questions regarding healthcare AI transparency.

|  |  |
| --- | --- |
|  | **N (%)** |
| **Importance of being informed by care team about every AI use in your care** |  |
| Very important/Important | 28 (66.7) |
| Neutral/Unimportant/Very unimportant | 14 (33.3) |
| **Importance of participating in decision-making regarding use of AI in your care** |  |
| Very important/Important | 28 (66.7) |
| Neutral/Unimportant/Very unimportant | 14 (33.3) |
| **Importance of getting information about how AI is used in your care** |  |
| Very important/Important | 30 (71.4) |
| Neutral/Unimportant/Very unimportant | 12 (28.6) |
| **Do you think labels for AI software are needed?** |  |
| Yes | 33 (78.6) |
| No | 9 (21.4) |
| **Importance of provider explaining how they made a decision about your care based on AI result** |  |
| Very important/Important | 31 (73.8) |
| Neutral/Unimportant/Very unimportant | 11 (26.2) |

### Information about the AI tool, oversight, and impact on care experience influence trust

Participants expressed information needs that generally fell into one of several major and sub-themes including information about the tool, oversight mechanisms, and impact on care as illustrated in a thematic map in Figure 1.

**Figure 1.**
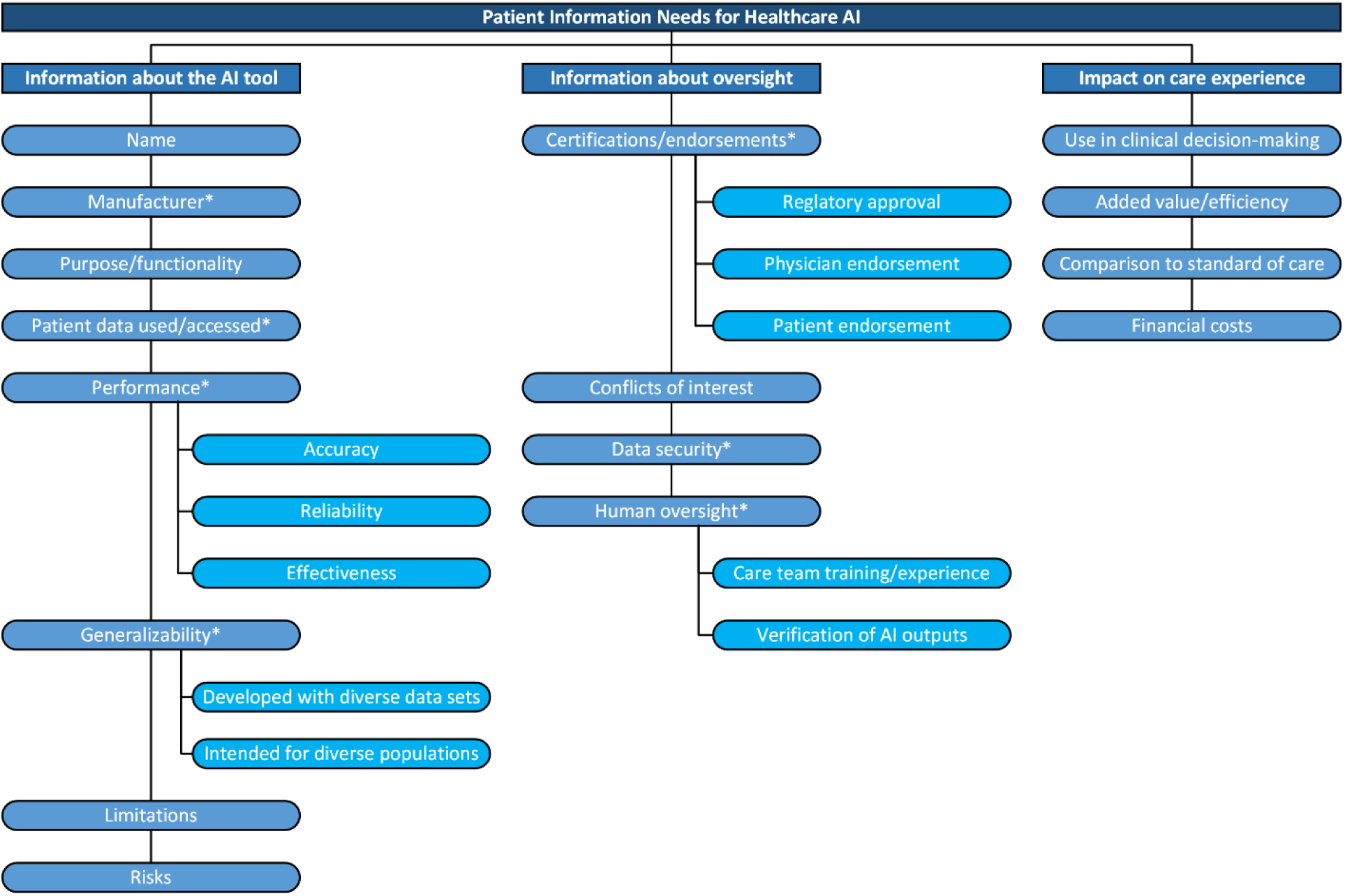
Thematic map of patient information needs for healthcare AI. High-level categories of information needs identified from inductive analysis include information about the AI tool, information about oversight, and the impact on the care experience. Information factors are enumerated for each category. Asterisks indicate factors that mediate trust.

### Information about the AI tool

Participants wanted to know the general characteristics of AI tools used in their care such as its name, function, and purpose. Participants also wanted to know who manufactured the tool, which influenced their trust of a given system due to perceptions of reliability.

> *I would want to know the company that produced the software in the smart watch and the company that produced the smart watch. Their reliability tells more about the accuracy of the software. (FG3)*

Patients also sought more evidence-oriented information such as the training data used, past performance, and its generalizability. Discussions of AI performance by patients concentrated on tool accuracy, its proven effectiveness, and reliability. These factors impacted perceptions of trust and effectiveness and were relevant to concerns about algorithmic bias and errors.

> *I would want to know what studies were done using the particular AI being used in my healthcare - and how reliable, effective, and what outcomes/impact on care it had. (FG3)*

> *In order to trust the results, I guess I’d want to know that people like me have been represented in the tests, clinical trials, etc. (FG2)*

Participants also wanted to be informed about a tool’s limitations and potential risks.

> *I would want to know about any limitations needed while using it, which the provider would usually review with me anyway. Trust results if verified as correct by provider. (FG1)*

### Information about oversight

Patients wanted to know what oversight procedures were in place for any AI tools used in their care which was also a factor influencing trust. Effective oversight was described as a distributed and layered process undertaken by regulatory bodies, care teams, and even other patients.

> *I would imagine a board with medical providers and patient representatives setting rules and procedures for all kinds of situations and monitoring compliance. (FG1)*

> *I would feel comfortable, knowing fully well it’s been approved by FDA and other relevant agencies governing it’s [sic] usage. (FG3)*

Additionally, a recurring theme was patients’ preference for their care teams to review and confirm the insights from AI systems used in their care, leading to greater trust.

> *Trust in the AI will come when my healthcare team reviews the data collected and agrees with the AI generated recommendations. (FG1)*

Lastly, participants felt strongly that there should be transparent and reliable oversight mechanisms in place for the use and storage of their data. They wanted to know what data stewardship practices were in place with particular concern for whether their data might be shared or sold, which influenced their trust in these systems.

> *I see this as a regulatory concern. I would think that AI presents a big logistical challenge with regard to sharing patient data culled from medical records. Privacy and confidentiality need to somehow be maintained. (FG1)*

### Impact on care experience

Participants wanted to know precisely how AI tools would be used in their care such as how AI would factor into clinical decision making, the added value AI provides, and how care involving AI compared to standard of care procedures.

> *I would appreciate transparent and clear communication about the role and impact of utilizing AI in that healthcare service setting. (FG3)*

> *I would want to know how it is done and comparable to an in-office test, something that was told to me before I actually got the warning to trust that it is accurate. (FG2)*

Participants also desired information on any financial costs associated with the use of AI in their care.

> *I would want to know how much this [AI] is costing me. Especially if this is something you would usually have as a follow up with a medical professional. (FG2)*

### Preference for model documentation and information delivery via labels and other mechanisms

Most participants were receptive to the idea of labeling AI tools. This was reinforced by the multiple-choice question data, with 78.6% (n=33) participants responding that labels would be necessary for the use of AI tools in their care. Patients also emphasized that labels should be designed with an emphasis on interpretability for patients.

> *Labels for AI software are a great idea, in principle. They need to be understandable to the layman. (FG1)*

However, some participants challenged the idea of an informational label suggesting that they might contribute to misunderstanding. Labels could potentially create confusion or fail to adequately communicate complex information about AI models.

> *I think the struggle with this is the communication of what these labels mean. It is not common knowledge of everything that is described on nutrition labels, and I think it would be similar and could lead to confusion in the health care setting further. (FG2)*

> *Labels are good, but might oversimplify the precision and huge amount of data, processing and nuances that go into any AI technology. (FG3)*

Patients also suggested other mechanisms for providing information on AI tools, including leveraging consent forms, informational videos, visit summaries, or discussions with their care team.

> *I think that any information that could be included on a label is best contained in a consent form signed with your doctor for treatment of a specific condition. Since different AI software would be used for different scenarios, the label would vary accordingly. It needs to be linked to the specific condition it is being used to treat. (FG1)*

In addition to specifying *how* they wanted information delivered, participants also commented on *who* they would like to receive information from. Participants often stated wanting to be informed by their treating physician or other member of the care team.

> *Practically I would love the information about this AI from an experienced practitioner. I would prefer the information be explained to me by an expert either directly or through emails. (FG1)*

Patients also discussed the appropriate timing for receiving information, with many participants wanting to be informed about the AI tool prior to or during its use for tools that are higher risk or are addressing more immediate concerns. Participants also indicated information about the AI tool used could be included in post-visit documentation such as an after visit summary.

> *I would want to be notified on the spot directly from my Doctor, I would like the info presented to me both on paper in an after visit summary. I would also like it to be verbalized to me by that same Doctor. (FG2)*

### Importance of disclosure of AI use when critical to provider decision making or care provision

On the multiple-choice survey questions, 66.7% (n=28) participants indicated that it was very important or important to be informed by their care teams about every use of AI in their care, and 71.4% (n=30) participants responded that it was very important or important to receive information about how AI is used in their care. Focus group responses also indicated, most patients wanted to be informed about AI use to understand the impact on their care and how it might factor into a care team’s decision.

> *AI is a tool that can be used by the team and I would like to know how it is used. Since AI is a relatively new technology it is important that the patient is informed in order to build trust in AI. (FG1)*

Some patients noted that the context of AI use influenced their perceived importance of its disclosure; patients wanted AI to be disclosed if it played a critical role in their care (e.g., surgical procedures, diagnoses, monitoring), but were ok with not being informed about less consequential uses (e.g., scheduling, notifications).

> *In small things like appointment scheduling, I think it is obvious it is AI and I do not need to be informed for it. In bigger things like device monitoring I would probably like to know. I think knowing for me is more so just because if something goes wrong, I will know who to contact or what to do. (FG3)*

Participants were mixed on being alerted to AI findings and, if so, how. Some welcomed being provided with all data gathered by AI, while others only wanted to be alerted by their doctor if AI detected something serious, warranting treatment. Some worried that notifications about minor findings could cause undue alarm.

> *I think that a lot of caution is needed in presenting AI analysis results. They could cause a lot of unnecessary stress to individuals involved. I also think this need[s] ethical considerations. What if an AI analysis predicts a non-curable, terminal illness? What are the ethical considerations for that kind of analysis and information. (FG1)*

### Variability in patient perceptions of consent to use AI

Participants expressed mixed opinions regarding the need to provide consent for AI to analyze their data. Some thought their data was likely already being used and that consent was already covered under agreement to be treated.

> *I could not care less. I almost have some expectation that it [AI] will be utilized without my knowledge or consent. I don’t see the need to make this any different than a doctor using any computer system to assist them. (FG2)*

Others valued consent but did not see the need for it to be provided each time AI was used. Some patients thought a broad consent would suffice; others wanted to provide consent for particular uses. For instance, participants were more concerned about consenting when their personal information might be shared with third-parties and less concerned when used by their care team for treatment.

> *I would feel uneasy about this unless it was disclosed and I consented to the continuous monitoring. Once consented, similar to the waiver for all healthcare data collected at [Clinic] to be open for research purposes, I found this honest and innovative that my data may help others in the future. (FG2)*

Finally, some participants wanted to provide consent every time AI was analyzing their data.

> *I would have to consent to any of this. It sounds like overkill. I would be concerned about the security of the information. (FG1)*

### Variability in patients’ preferences for deciding whether AI is used in their care

66.7% (n=28) participants indicated that it was very important or important to be involved decision-making regarding the use of AI in their care. Furthermore, 73.8% (n=31) responded that it was important to have a provider explain how a decision was made based on an AI result. Patients expressed varied opinions on needing to be involved in decision-making concerning healthcare AI. Many felt they should be involved in the decision to use AI just as they would want to actively make decisions about other aspects of their treatment.

> *My decision must be required before I can allow a machine work on me even if it’s 100% accurate in its dealings and more accurate than humans. (FG3)*

Other participants envisioned leaning on the expertise of their care team, but still maintaining authority in deciding whether AI would be used.

> *If the provider is supposed to be an expert, I expect them to give me their opinion about what’s best. Of course, as a patient, I would always have to right to yay or nay its use. That said, I always anticipate being a partner in my healthcare decisions with any providers. (FG2)*

Some patients were comfortable deferring to their care team’s judgment, suggesting that they would have more expertise to make an informed decision. However, informing patients about the use of AI was still desired.

> *I’m not sure I’m qualified enough to make an informed decision about its use. I would like to know if and how it’s being used but I would leave the decision up to the care team. (FG1)*

### Mostly favorable perceptions of physicians using AI

Participants generally expressed that their opinion of their provider would not change if their provider used AI in their practice. Furthermore, others trusted their physician’s judgment on when AI might be a helpful tool based on the physician’s clinical knowledge.

> *It [AI] would not impact my trust for the experienced physician at all because ultimately, I trust that physicians use tools and experience to come up with the diagnosis. The AI is there as a technologically enhanced tool. (FG1)*

Many viewed the use of AI as a positive sign that their physician was keeping up to date with new technology and using it in conjunction with their clinical expertise to provide care and make recommendations.

> *It wouldn’t change my opinion in a bad way but might in a good way. For example, I love to read and am often surprised that I might have read something that a clinician has not. It makes me think that they don’t try hard enough to keep up with new research. If I discovered that a provider was using AI software, I think I’d think more highly of them, depending on the purpose of the software. (FG2)*

The extent to which participants had cultivated relationships with their physicians over significant periods of time contributed to this level of trust, irrespective of the tools used.

> *It would not change my opinion of my healthcare provider. How I feel about him/her is based on personal interaction, knowledge of my medical history, and the trust that has been built up over the years. (FG1)*

In circumstances where a physician disagreed with an AI recommendation, most participants would not view the physician negatively. Some patients would even view their physician more positively for not deferring to the AI recommendation.

> *I would believe that the provider is thinking critically about the results of the AI’s interpretation of the data it was given, and as long as the explanation and rationale is given plausibly and logically, I would be reassured by the disagreement. If it kept happening though repeatedly in a pattern, it would concern me that something systemically, on either (or both) [the] part of the prescriber or AI, is wrong. (FG3)*

However, some participants would want reassurance if their physician disagreed with the AI’s recommendation and might seek out a second opinion from another physician.

> *I usually do some research before even seeing a provider so I have a notion of what they’ll say and even prescribe. If a doctor disagrees with an AI’s result but still aligns with what I’ve found on my own, that would be helpful for me but I think I’d still have to seek a second opinion and let them know why I was doing that. (FG2)*

## Discussion

The present study is among the first to systematically identify information needs and factors that facilitate trust in AI from the perspective of patients. Our findings add to the growing body of work in this area by expanding, affirming, and characterizing patient information needs and trust factors identified from other specialty areas.^23^ Furthermore, our work expands on past research demonstrating transparency is important for fostering trust in the use of AI systems in healthcare and physicians who chose to use AI.^41^ While our findings are grounded in clinical vignettes describing AI in cardiovascular care, it is worth noting that patients often generalized their responses to broad uses of AI in their care in addition to responding to specific vignette-based questions.

While there is consensus around the need for transparency, the best way to deliver key information that promotes transparency remains less clear. Many, both internal and external to the healthcare sector, have proposed model documentation in the form of AI labeling and sharing of associated data sets.^19, 20, 42, 43^ Dynamic AI labeling, in particular, has been argued to be a critical part of supporting overall regulatory approaches for healthcare AI.^44^ Our results show that while patients tend to think labeling could improve transparency, labels may not always be the best or most comprehensive approach for communicating this information. Labels instead might serve as an entry point to a broader ecosystem of information that patients can engage with according to their needs and preferences.

Patients have concerns about labels being confusing and inaccessible and therefore may prefer alternative ways of delivering this information (e.g., consent forms). This finding highlights the importance of incorporating diverse patient perspectives when designing strategies to disseminate information about AI tools and tailoring these strategies according to patient needs, preferences, and values. For instance, progressive disclosure could be used as an approach to allow patients to decide the depth of information they would like to review about AI tools used in their care, and materials could then be created accordingly (e.g., label with brief facts, patient handouts with more in depth data and information).^45^ Visual and interactive aspects of model documentation could also be considered in support of patient needs, and further highlights the relevance of user interactions with AI systems when designing for transparency.^46^ Additional research is needed to identify what mechanisms are most effective at delivering information and calibrating trust in AI systems and the care teams that employ them, and how to tailor delivery mechanisms to accommodate diverse patient preferences and clinical scenarios.

Our findings suggest that transparency for patients encompasses more than information availability. Patients often express a preference for being an active decision-maker in their care and use of AI in it, albeit to varying degrees. The presence of healthcare AI in clinical settings may prompt additional conversations between patients and their care teams regardless of whether labels or other delivery mechanisms were used to communicate the information. While ubiquitously stated in ethical frameworks and guidelines, transparency lacks consensus and is often complicated by the audience AI systems need to be transparent to.^2, 47^

Moreover, our findings suggest physicians play a pivotal role in facilitating AI transparency and mediating patient trust. Our findings suggest that patients will often look to their physician(s)’ judgment on whether AI tools are safe and effective in their care, consistent with findings from other patient-centered research.^48^ These insights emphasize a need to further understand how physicians develop trust in AI tools and how that trust may then be shared with patients.^41, 49^ Furthermore, there are numerous implications for physicians in the adoption of AI tools, such as training, disclosure, consent, and liability that are relevant to how associated risks may be communicated to patients.^12, 50-52^ These additional responsibilities for physicians and healthcare institutions should be evaluated as regulations are considered for AI systems.

Developing AI transparency solutions for patients fits into a broader range of strategies for enhancing overall transparency of these systems in healthcare, including for clinical and technical stakeholders. This manuscript focuses on the information and transparency needs of patients. This focus is particularly important given the dearth of research with patient populations, even though their personal data will likely be used for the development of healthcare AI systems, and they will be on the receiving end of clinical decisions driven by these tools.^53^ By including patient perspectives in ongoing transparency work, there are opportunities to support health equity and foster trust in healthcare AI tools.^54^ In coordination with technical approaches, ethical frameworks, and regulatory guidelines, there is the ability to strengthen healthcare AI transparency for all stakeholders in accordance with their precise information needs.^17^

Our study has several limitations. Conducting focus groups in an online asynchronous format constrains the moderators’ ability to ask follow-up and clarifying questions.^30^ Some follow-up questions did not receive a response, and sometimes there was a time delay in correcting misunderstandings to probes. Participants were also limited to those who had sufficient broadband access and technological literacy to participate in an online hosted discussion.^55^ Additionally, our findings might have been impacted by selection bias, limiting their generalizability. While ResearchMatch helped facilitate recruitment beyond a single healthcare institution and local geographic region, as an online health research-oriented database, the pool of potential participants may have possessed greater health and technological literacy than that of the general populace. Future research should further engage underrepresented populations and communities through participatory research methods to examine whether and how AI information and transparency needs differ across patient demographics, with the goal to inform AI information dissemination strategies and policies that are better tailored to varying community needs.

Participants were prompted to respond to uses of AI in healthcare broadly and more specifically through clinical vignettes based on four AI applications in cardiovascular care. However, there is immense variety in healthcare AI between numerous clinical specialties and ML methods. Thus, further research evaluating a broader range of technologies and their use in clinical specialties is necessary to better understand the nuances of information needs as they relate to specific AI applications and care circumstances. While our findings suggest that there are a set of information needs relevant for patients, our analysis is limited in its ability to generalize and characterize the relative importance of these information needs. Additional research with larger study samples is needed to better understand how these factors might be prioritized by patients for potential transparency solutions.

## Conclusion

As healthcare stakeholders grapple with the adoption of AI into clinical settings, consideration for patient information needs will be vital for calibration of trust and determinations about safe and effective use of these tools. The present study outlines several of those needs along with key patient perspectives on information delivery, decision-making involvement, and physician AI use. While some of these factors overlap with transparency solutions outlined for technical and clinical experts, insights from patients can aid the expansion of these approaches to better support patient-centered adoption of healthcare AI.

## Data Availability

All data produced in the present study are available upon reasonable request to the authors.

## Acknowledgements

Not applicable

## Author contributions

AMS and SAM contributed to data curation, formal analysis, investigation, and writing. XZ contributed to the conceptualization, methodology, data curation, formal analysis, investigation, and writing. JLR contributed to the conceptualization, methodology, and writing. JEM contributed to the conceptualization, funding acquisition, and writing. BAB contributed to the conceptualization, funding acquisition, methodology, supervision, data curation, formal analysis, investigation, and writing.

## Ethical approval

The Mayo Clinic Institutional Review Board approved this study (protocol # 21-012302). All participants consented verbally prior to participate in each focus group and to the distribution of their de-identified information in accordance in accordance with IRB approvals.

